# Evaluating the sensitivity of heart rate variability fractal correlation properties to training load variations: Implications for monitoring training readiness and durability

**DOI:** 10.64898/2026.05.27.26354281

**Authors:** Cody R. van Rassel, Markus Rummel, Martin J. MacInnis

## Abstract

This study examined the utility of HRV detrended fluctuation analysis alpha-1 (DFAα1) to assess readiness-to-train and exercise durability under varying acute training loads. Nineteen trained cyclists completed two 20-minute time-trials (TT) under rested and fatigued conditions. DFAα1 was measured during a standardized warm-up (WU), 20-min TT, and standardized cool-down (CD). Power output (PO) and DFAα1 responses were compared across conditions, and associations with performance and fitness (W·kg^−1^) were examined. DFAα1 values declined with increasing WU and CD exercise intensity (p<0.001) and were significantly attenuated following the 20-min TT (p<0.001). While DFAα1 profiles did not differ significantly between rested and fatigued conditions, lower pre-TT DFAα1 was associated with reduced TT performance (p=0.022; r=0.55), suggesting relevance to training readiness. Additionally, an 18% decline in DFAα1 between 10- and 20-min during the TT (p=0.031), and lower post-TT values at matched intensities were observed (p<0.001), indicating physiological perturbation from the 20-min TT. Fitter participants exhibited lower DFAα1 values during the 20-min TT (p<0.001; r=−0.77), suggesting a greater capacity to sustain physiological stress. While DFAα1 is responsive to exercise intensity and stress, offering potential to assess training readiness and durability, more robust fatigue protocols are needed to validate DFAα1 as training load monitoring tool.

## Introduction

Periods of intensified training are essential in endurance sports, promoting performance gains through functional overreaching when balanced with sufficient recovery (Aubry et al., 2014). Excessive training without adequate rest; however, can lead to non-functional overreaching or overtraining, resulting in prolonged performance decline (Le Meur et al., 2013; Meeusen et al., 2013). Aligning training strategies with fluctuations in physiological capacity may help optimize adaptations (Halson, 2014). In this context, training load has become a widely adopted concept, originally proposed as a framework to quantify overall training “dose” and predict adaptive responses (Banister et al., 1975). Despite the development of numerous training load models, most have not been validated as reliable indicators of training dose or performance (Passfield et al., 2022). Accordingly, there is a need for practical tools that can accurately quantify accumulated training stress in relation to exercise performance outcomes. Such methods could improve training strategies and reduce non-functional overreaching, injury, and illness (Halson, 2014).

One popular approach to monitoring training load is heart rate variability (HRV). Typically, non-exercising HRV indices like high frequency (HF) power and root mean square of successive differences (RMSSD) have been used to assess training readiness, recovery, and functional overreaching (Coates et al., 2018; Plews et al., 2013; Stanley et al., 2013). While promising, their interpretation is often contingent on testing conditions (e.g., posture, environment) and analysis method, raising concerns about their reliability for informing training decisions (Bellenger et al., 2016; Coates et al., 2018; Manresa-Rocamora et al., 2021). Assessing HRV in an exercising state, where physiological perturbations are much greater than at rest, may provide timely insights into training responses.

Recently, adaptations of non-linear dynamics and chaos theory have been applied to HRV with the aim of quantifying the complex regulation of cardiovascular activity influenced by neural and non-neural sources during exercise (Rogers & Gronwald, 2022). Unlike traditional time and frequency domain metrics, which provide limited insights under exercising conditions, non-linear HRV metrics are highly responsive (Rogers & Gronwald, 2022). Detrended fluctuation analysis (DFA), for example, is a non-linear technique used to estimate non-stationarities within time-series data. By fitting a power law to the average signal fluctuations across multiple time scales, the slope of the log-log plot between fluctuations and time scales provides the short-term scaling exponent, alpha-1 (DFAα1) (Peng et al., 1995).

DFAα1 may provide valuable insights into between-session and within-session physiological perturbations related to exercise training. During incremental exercise, DFAα1 responds dynamically to increasing exercise intensity, with values shifting from >1.0 at light-to-moderate intensities toward 0.75 and 0.5 near heavy and severe intensities, respectively, indicating a progressive loss of signal correlation (Rogers & Gronwald, 2022). DFAα1 is also sensitive to prolonged exercise, highlighting its potential value to capture cumulative training stress across various intensities and durations (Ajayi et al., 2025; Gronwald et al., 2021; van Rassel et al., 2024). For example, changes in DFAα1 at fixed relative intensities before and after a training session can estimate the amount of fatigue accumulated from ultra-endurance efforts (Rogers, Mourot, et al., 2021) and distinguish between varying levels of training stress (Ajayi et al., 2025; Schaffarczyk et al., 2022). The degree of DFAα1 signal attenuation during prolonged running has also been linked to reductions in lactate threshold evaluation (Nuuttila et al., 2024). These findings highlight the potential of DFAα1 to indicate altered states of training “readiness” and quantify physiological shifts or deterioration during prolonged exercise—a concept commonly referred to as exercise durability (Maunder et al., 2021). Despite this theoretical understanding, DFAα1 profiles—both between and within session—have not been systematically evaluated in relation to exercise performance outcomes, and it is unclear whether DFAα1 can indicate readiness-to-train, predict performance outcomes, or examine exercise durability.

The purpose of this investigation was to determine whether exercising DFAα1 measurements can distinguish between periods of intensified training and predict variations in performance outcomes. Using a standardized exercise protocol performed before and after a 20-min cycling time-trial (TT), in both simulated “rested” and “fatigued” conditions, we examined whether DFAα1 can serve as an indicator of readiness-to-train and durability under varying training loads. We hypothesized that distinct DFAα1 profiles would emerge that correlate with training load, exercise performance, and athlete fitness, thereby providing indications of training readiness and exercise durability.

## Materials and Methods

### Participants

Nineteen (3 female, 15 male) recreationally (n=7) and competitively trained (n=12) cyclists (mean [SD]; age = 36 [10] years, body mass = 77.0 [9.0] kg; height = 179.7 [9.3] cm) were recruited for exercise testing. Fourteen participants conducted their exercise testing in a laboratory at the University of Calgary. The remaining five participants completed the exercise testing sessions remotely. Over the 3 months prior to testing, participants reported performing endurance exercise 4.4 [1.7] days per week and 6.7 [4.3] hours per week (2.6 [1.4] days per week and 3.9 [2.8] hours per week cycling). Eleven participants reported training to compete in an endurance event.

Written informed consent was provided by each participant prior to completing the experimental procedures, which were approved by the University of Calgary Conjoint Health Research Ethics Board (REB22-0159) and performed in accordance with the Declaration of Helsinki. A Physical Activity Readiness Questionnaire (PAR-Q+) was completed to identify contraindications to exercise testing and to ensure that no medical conditions or injuries were present that could interfere with cardiorespiratory exercise responses.

### Experimental Design

Within a 14-day period, participants performed four separate exercise testing sessions on a cycle ergometer. Sessions consisted of a 4-min time-trial (TT), a “rested” 20-min TT, a virtual cycling race, and a “fatigued” 20-min TT. Participants were instructed to prepare for each exercise session as if it were a competitive race (i.e., arrive rested, hydrated, and fuelled for intense exercise), match their diets and caffeine use across sessions, refrain from consuming caffeine or eating within 2 hours prior to each session, and avoid drinking alcohol or engaging in exercise (beyond the study requirements) within 48 hours prior to their sessions. Music was not permitted during the testing procedures. Participants were asked to rate how well rested they felt prior to their exercise sessions on a −5 to +5 point scale—0 indicating average, and −5 and +5 indicating extremely unrested and well-rested, respectively. Rating of perceived exertion (RPE) was evaluated following each exercise session using the Borg RPE scale (6–20) (Borg, 1982).

### Equipment and Measurements

All exercise testing sessions were performed on a stationary cycle ergometer and conducted using the online cycling training program, Zwift (Zwift Inc., Long Beach, California, USA)—a virtual training platform that combines gaming with stationary cycling. All participant heartbeat RR-interval data were collected using a Polar H10 chest strap (Polar Electro Oy, Kempele, Finland) with a sampling rate of 1000Hz. Laboratory-based testing was conducted on a Garmin Tacx Neo Bike Smart Trainer (Garmin Ltd., Olathe, Kansas, USA), with RR-interval and PO data recorded via Bluetooth (Bluetooth SIG, Kirkland, Washington, USA) using a Forerunner 955 sport watch (Garmin Ltd.). Remote-based testing was performed using Wahoo Kickr (n=3; Wahoo Fitness, Atlanta, Georgia, USA), Wahoo Kickr Core (n=1), or Zwift Hub smart trainers (n=1; Zwift Inc.), with RR-interval and PO data recorded via Bluetooth using an Edge 530 (n=3) or Edge 1030 (n=2) cycling computer (Garmin Ltd.). With additional participant consent (REB20-0572), researchers accessed remote-based participant data, uploaded via secure third-party platforms (e.g., Garmin Connect), through the University of Calgary’s Wearable Technology Citizen Science Program web portal and Level-4 secure MySQL database.

### Exercise Protocols

All cycling TTs were performed in the morning before 12:00 pm, and at approximately the same time each day. The 4-min TT, “rested” 20-min TT, and virtual cycling race, were scheduled at least 48 hours apart. To create the “fatigued” condition, participants performed a virtual cycling race the evening (after 4 PM) prior to the fatigued 20-min TT. The 4-min TT was performed during the first session, but rested and fatigued TTs were performed in a randomized order.

For laboratory participants, functional threshold power (FTP) was estimated as 75% of the average 4-min TT PO achieved (MacInnis et al., 2019); however, for remote participants, FTP was self-reported using historical training data on the Zwift platform (Zwift Inc.) to avoid delays in testing. The self-reported FTP of remote participants (271 [58] W) was not different from the FTP predicted from their 20-min TT (i.e., 90% of rested 20-min TT PO; 270 [56]; p=0.861; n=5) (MacInnis et al., 2019).

### 4-min and 20-min Time-Trials

Participants were instructed to complete each TT at the greatest possible PO they could maintain for the prescribed duration and were permitted to change gears (real or simulated). Cycling PO and HR were hidden during the laboratory-based TTs and remote-based participants were instructed to hide these variables as well.

Five minutes before and after the rested and fatigued 20-min TTs, participants performed a standardized warm-up and cool-down, consisting of cycling at 60% (6 min), 70% (6 min), and 80% (3 min) of FTP (Figure 1). The same warm-up and cool-down workloads were used for both the rested and fatigued 20-min TT conditions. Remote participants used their personal equipment and Zwift profiles to complete the study protocols.

**Figure 1:**
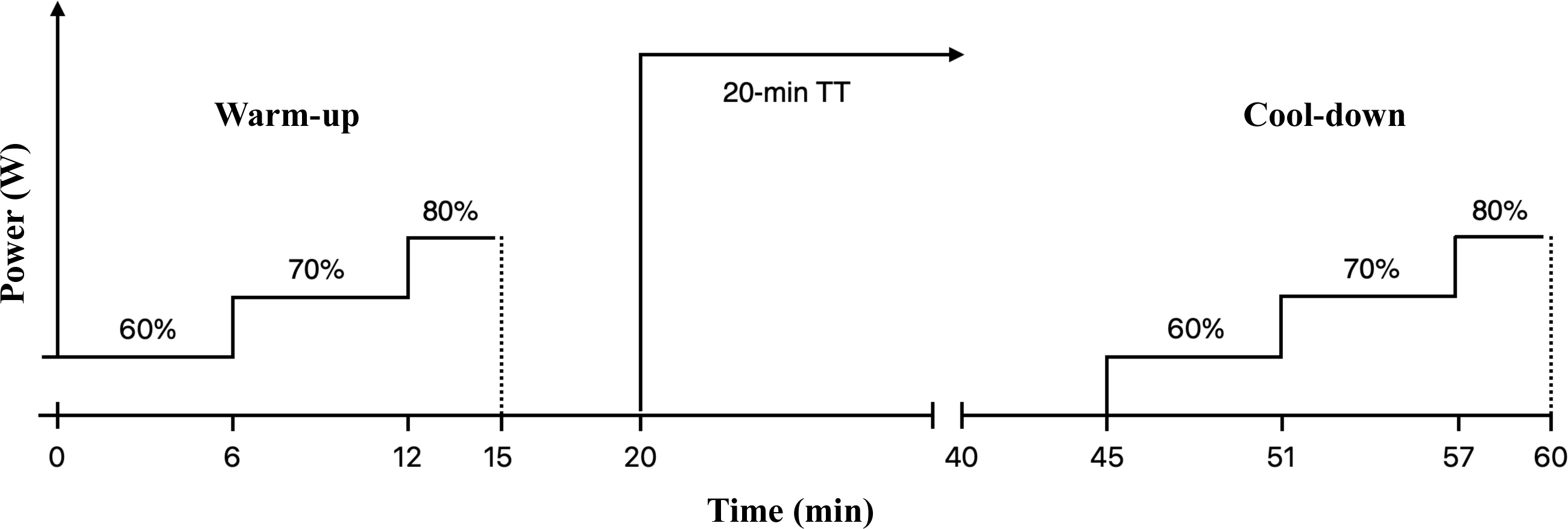
Schematic of the exercise testing protocol. Diagram illustrates the 20-min time-trial (TT) exercise protocol for both the “rested” and “fatigued” conditions. Each 20-min TT began with a 15-min structured warm-up (WU) consisting of 6-min, 6-min, and 3-min of constant-load cycling at 60, 70, and 80% of estimated functional threshold power (FTP), respectively. Participants passively cycled for 5-min before performing the 20-min TT, which was followed by 5-min of passive cycling and a 15-min structured cool-down (CD), with workloads and durations identical to the WU.

### Virtual Cycling Race

The virtual cycling races were selected from a variety of available online Zwift races where the estimated time to completion was between 30-45 minutes. Virtual race difficulty was selected based on each participants estimated FTP (i.e., W·kg^−1^), ensuring that each race offered a substantial challenge among cyclists of similar fitness levels. Participants were instructed to complete the virtual race with a maximal effort.

### Data Analysis Data Processing

Heartbeat RR-interval data were imported into Kubios HRV Premium Software version 3.5.0 (Kubios Oy, Kuopio, Finland) for preprocessing prior to analysis. RR-interval detrending was set to “smoothness priors” (Lambda = 500), automatic noise detection set to “medium,” and beat correction threshold set to “automatic.” RR-interval data files with noise or beat corrections that exceeded 5% were excluded from subsequent analysis (Rogers, Giles, et al., 2021). Monofractal DFAα1 was calculated using a linear fit of the double log RMS fluctuation in relation to the different window sizes, with window width set to 4 ≤ n ≤ 16 RR-intervals (Peng et al., 1995). Using a modified Python script (NONAN, 2023), DFAα1 was calculated from the processed RR-interval data files using 2 min of RR-interval data (Hautala et al., 2003) and re-calculated every 5 s throughout the trials.

For final analyses, the average DFAα1 corresponding to mins 5-6, 11-12, and 14-15 (i.e., 60, 70, and 80% of FTP) of the warm-up and cool-down were calculated for both the rested and fatigued conditions. Additionally, the overall average DFAα1 across the entire warm-up and cool-down was determined for each condition. For the 20-min TT, the average DFAα1 corresponding to mins 9-10 and 19-20 were calculated for both the rested and fatigued condition, along with the overall average DFAα1 during the 20-min TTs. Participant fitness was calculated as the average PO achieved during the “rested” 20-min TT relative to body mass (i.e., W·kg^−1^).

### Statistical Analysis

Paired Student’s *t* tests were conducted to compare pairs of continuous variables, and Wilcoxon signed-rank tests were used for ordinal variables. Effect sizes for paired continuous variables were calculated using Cohen’s d (Cohen, 2013). Pearson’s correlation coefficients assessed the level of association. A three-way repeated measures ANOVA examined the effect of time-point (pre [warm-up] and post [cool-down] 20-min TT), condition (rested and fatigued), and intensity (i.e., 60, 70, and 80% of FTP) on DFAα1. A two-way repeated measures ANOVA examined the effect of duration (i.e., 10-min and 20-min) and condition (rested and fatigued) on 20-min TT DFAα1. A repeated measures ANCOVA examined the effect of condition on average DFAα1 during the 20-min TT. For the ANCOVA, the within-subject factor was condition and the covariate was mean PO. All assumptions for ANCOVA were checked and met, allowing us to remove the condition × PO interaction term from the final analysis. Estimated marginal means were calculated for each condition at the covariate value of the sample mean PO. ANOVA and ANCOVA effect sizes were assessed using partial eta squared (η^2^_p_) (Lakens, 2013). Data are reported as mean [standard deviation (SD)], with statistical significance set at an α level of <0.05. Where appropriate, Bonferroni post hoc tests were used. Analyses were performed using SPSS (version 26, IBM, Armonk, NY, USA), and data visualization was performed with Prism (version 10.3.1 for macOS, GraphPad Software, San Diego, CA, USA). Python programming language (version 3.7.7) was used to calculate DFAα1.

## Results

Participants with RR-interval noise or beat corrections >5% in the condition of interest were removed from subsequent analyses including two data sets from the warm-up (n=17), four data sets from the 20-min TT (n=15), and four data sets from the cool-down (n=15). All other parameters and measures included data from all 19 participants. Virtual cycling race durations were 35.3 [6.9] min. On average, 15.4 [2.0] h of time elapsed between the virtual cycling race and the fatigued 20-min TT. Average cycling PO was 3.4% lower during the fatigued (222 [50] W) compared to the rested 20-min TT (231 [50]; p=0.018; d=0.60; Figure 3). Participants reported feeling less rested before the fatigued 20-min TT (−0.3 [2.0] vs. 1.4 [2.0]; p=0.011), but post exercise RPE was not different between conditions (17.9 [1.9] vs. 18.1 [1.4]; p=0.331).

### Warm-Up and Cool-Down Analysis

Results from the three-way repeated measures ANOVA revealed main effects for time-point (F(1,14)=35.81, p<0.001, η^2^_p_=0.719) and intensity (F(1.43, 19.97)=42.80, p<0.001, η^2^_p_=0.754) on DFAα1 measurements, but the main effect of condition was not significant (F(1, 14)=1.40, p=0.257, η^2^_p_=0.091; Figure 2). Post hoc pairwise comparisons revealed that DFAα1 was significantly lower during the cool-down compared to the warm-up at each intensity level (p<0.001), regardless of condition, and was significantly different between all pairs of intensities during both the warm-up and cool-down (p<0.001 for all pairwise comparisons; Figure 2). The condition × time-point interaction (F(1, 14) = 3.36, p=0.088, η^2^_p_=0.194) and the time-point × intensity interaction (F(2, 28)=3.13, p=0.059, η^2^_p_=0.183) approached, but did not reach, statistical significance. The condition × intensity interaction was not significant (F(1.39, 19.48)=0.28, p=0.681, η^2^_p_=0.019; Figure 2), and the three-way condition × time-point × intensity interaction was also not significant (F(2, 28)=0.06, p=0.947, η^2^_p_=0.004; Figure 2).

**Figure 2:**
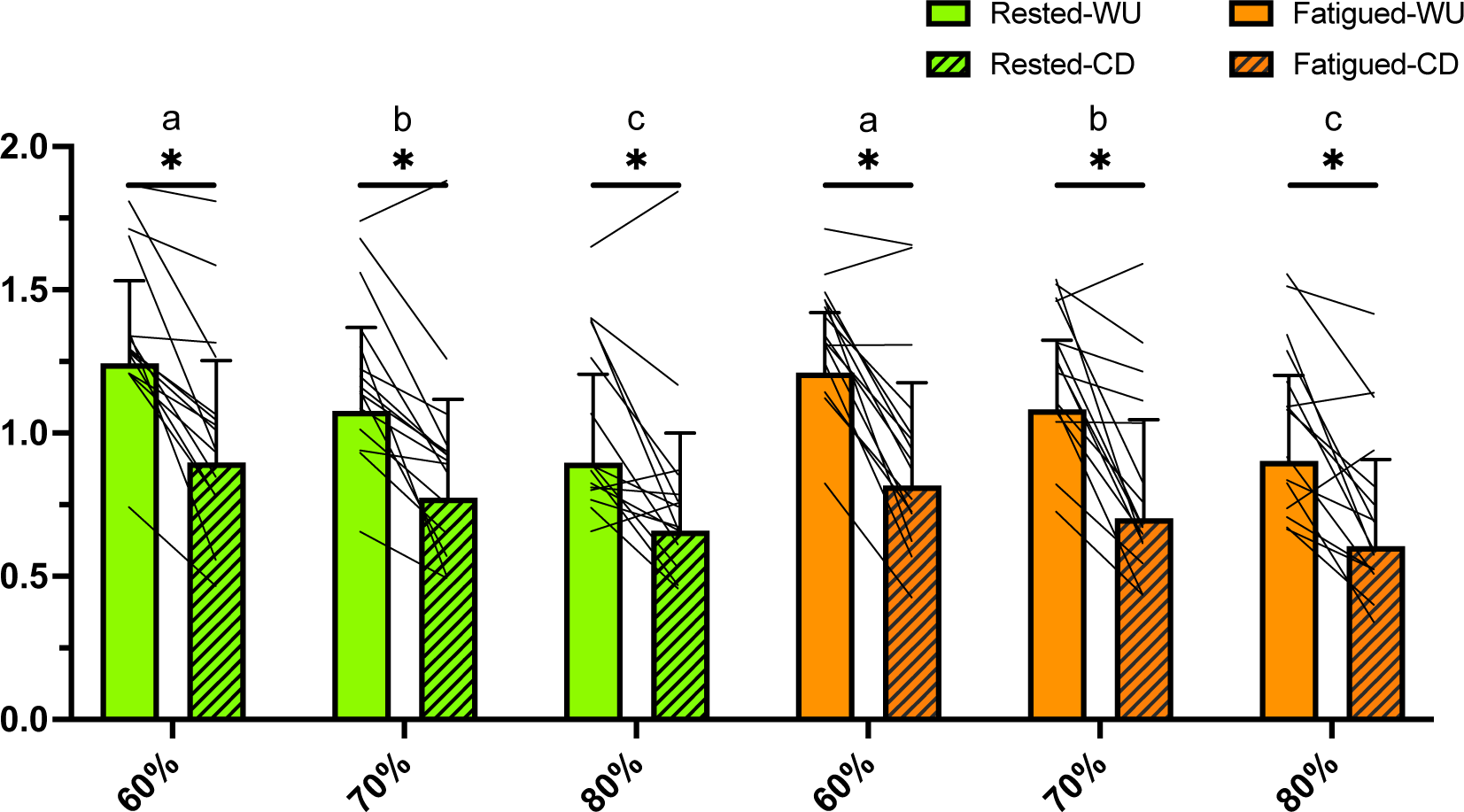
DFAα1 values across two time points (warm-up [WU] and cool-down [CD]), three intensities (60, 70, and 80% of functional threshold power [FTP]), and two conditions (rested and fatigued). Solid bars represent WU; hatched bars represent CD. Green = rested, orange = fatigued. Connecting lines represent within-subject responses. Significant main effects were observed for time point (p<0.001) and intensity (p<0.001), but not for condition (p=0.257). Interactions between condition and time point (p=0.088) and between time point and intensity (p=0.059) approached, but did not reach, statistical significance. Asterisks (*) denote significant differences between WU and CD at the same intensity level (p<0.001). Intensities that do not share a letter (a, b, c) are significantly different (p<0.001). N=15.

### 20-min Time-Trial Analysis

A significant main effect of duration (F(1, 14)=5.79, p=0.031, η^2^_p_=0.292) was detected for TT DFAα1, with the 20-min mean (0.47 [0.16]) 18% lower than the 10-min (0.58 [0.26]; Figure 3). On average, DFAα1 decreased by 11% and 24% between 10- and 20-min in the rested and fatigued conditions, respectively; however, the main effect of condition (F(1, 14)=3.89, p=0.069, η^2^_p_=0.217) and duration × condition interaction effect (F(1, 14)=2.387, p=0.145, η^2^_p_=0.146; Figure 3) were not significant. Average DFAα1 measurements during the full 20 min were not different between rested (0.57 [0.15]) and fatigued (0.62 [0.22], p=0.192, d=0.16; Figure 3) conditions. The mean PO was a significant covariate for DFAα1 when an ANCOVA was performed, with higher PO associated with lower DFAα1 values (F(1,27)=22.035, p<0.001, η^2^_p_=0.449); however, after adjusting for this covariate, there was still no significant main effect of condition for average DFAα1 during the 20-min TT (F(1,27)=0.156; p=0.696 η^2^_p_=0.006), with estimated marginal means of DFAα1 of 0.61 [0.14] for the fatigued condition and 0.59 [0.14] for the rested condition at the sample mean PO (240 W; Figure 3).

**Figure 3.**
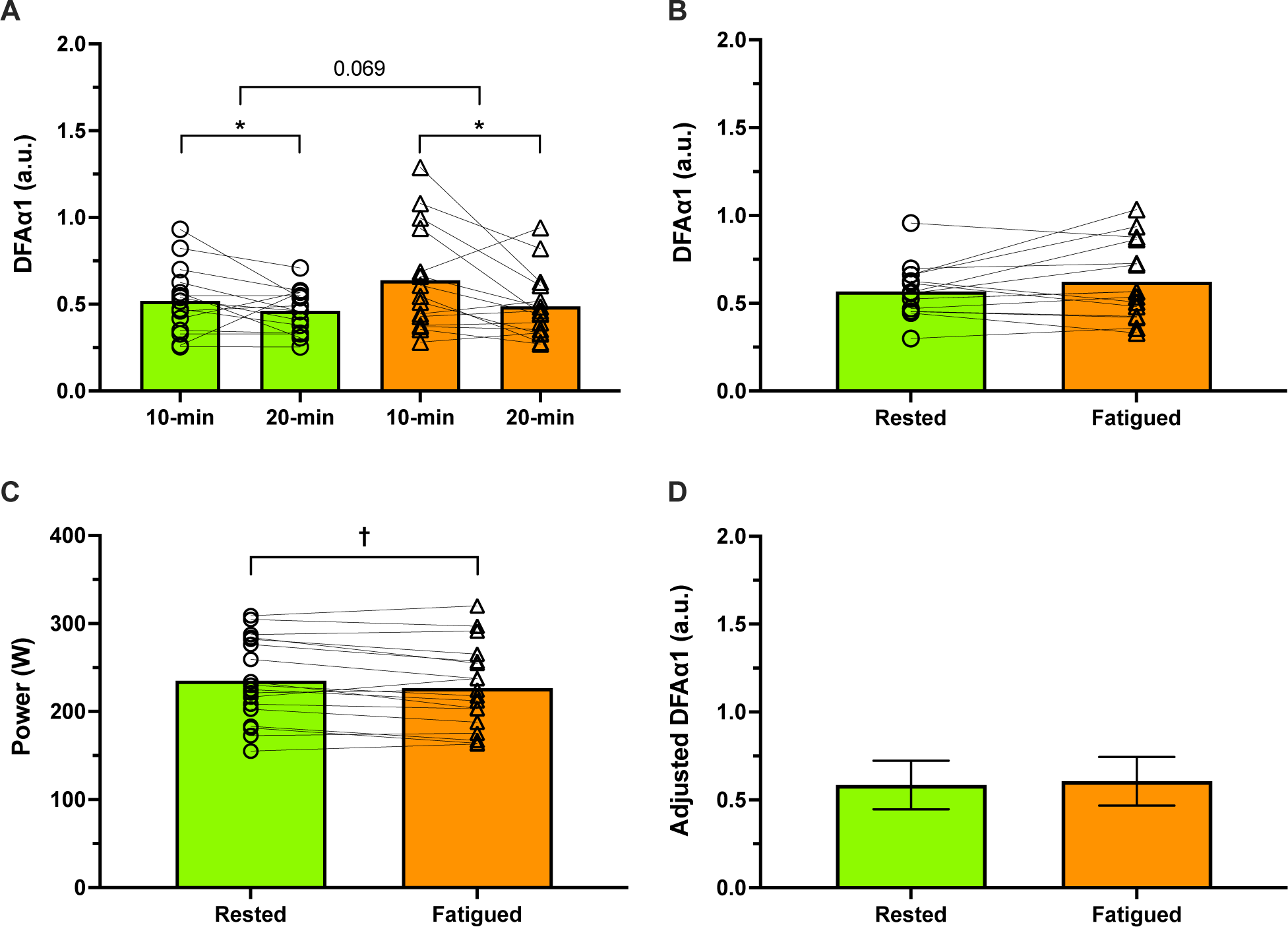
DFAα1 responses during the 20-min time-trials in rested (green) and fatigued (orange) conditions. Panel A presents the DFAα1 responses corresponding to the 10th and 20th minute of the time-trials; panel B presents the average DFAα1 measurements during the trials; panel C presents the average 20-min time-trial power output; and panel D presents the estimated marginal means (±SD) for DFAα1 in each condition, adjusted for mean power output (240 W) as the covariate in the ANCOVA analysis. P-values used to indicate trending main effect of condition. Asterisk (*) indicates significant difference between duration post-hoc comparisons (p<0.05). Dagger (†) indicates significant difference between conditions (p<0.05). n=15 for all panels.

### DFAα1 and Performance

A moderate positive association was detected between the difference in 20-min TT PO and the difference in warm-up DFAα1 between conditions (i.e., rested and fatigued) (Figure 4). These results indicate that relatively higher warm-up DFAα1 values were associated with relatively higher 20-min TT POs, independent of condition.

**Figure 4.**
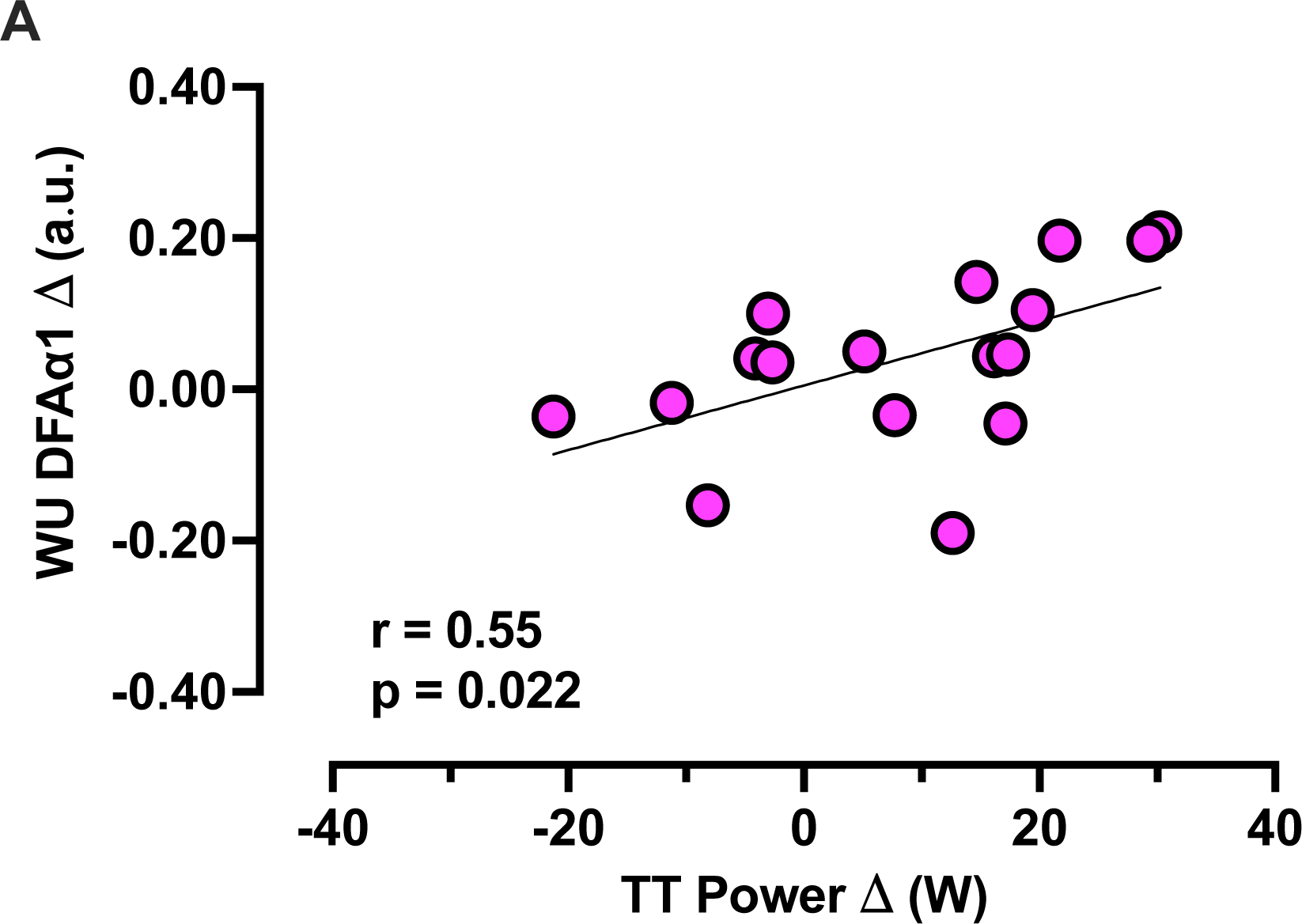
The relationship between the difference in 20-min time trial (TT) power output and the difference in warm-up (WU)-DFAα1 between rested and fatigued conditions. The regression line, Pearson correlation coefficient, and p values are provided. n=17.

### DFAα1 and Fitness

Average DFAα1 during the rested 20-min TT exhibited a moderate negative association with participant fitness, and average DFAα1 during the fatigued 20-min TT exhibited a strong negative association with participant fitness (Figure 5). These results indicate that fitter participants tended to exhibit lower average DFAα1 during 20-min of intense exercise.

**Figure 5.**
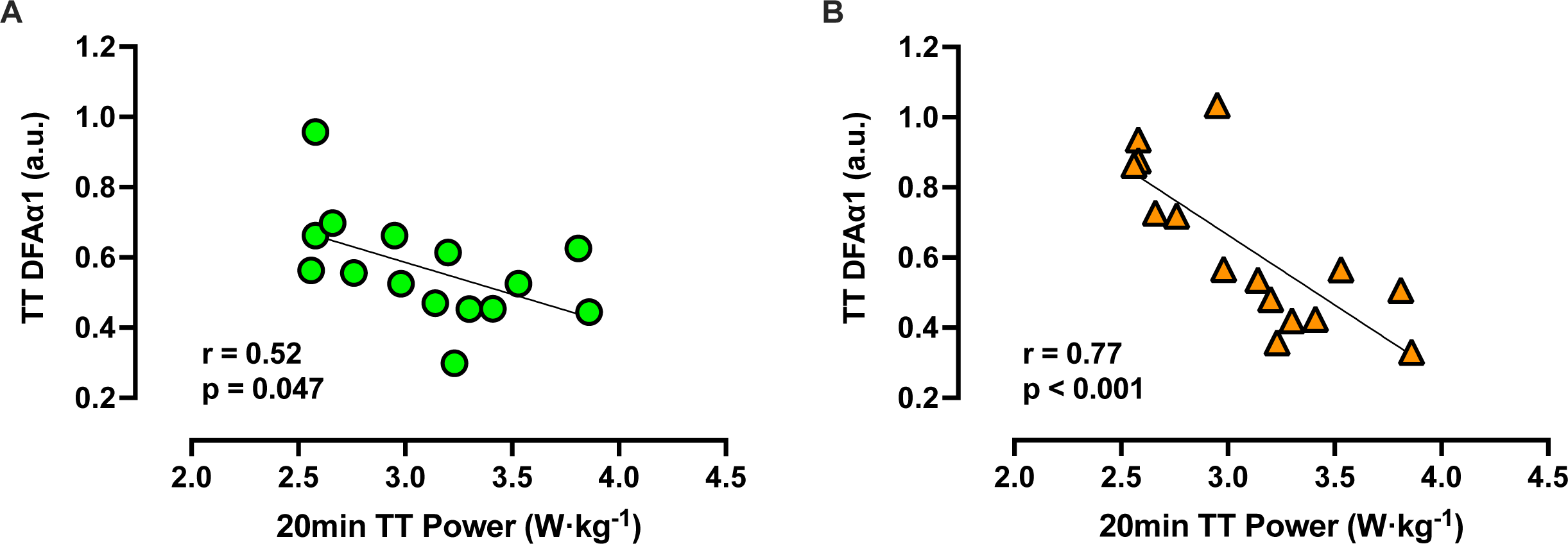
Relationship between the average DFAα1 during the rested and fatigued 20-min time-trials (TT) and participant fitness. Fitness was determined as the average rested 20-min TT power output (PO) relative to body mass. Regression lines, Pearson correlation coefficients, and p values are provided for each panel. n=15 for both panels.

### DFAα1 and Work Performed

In the rested condition, no significant association was observed between changes in DFAα1 from the warm-up to cool-down and total work performed during the 20min TT (Figure 6). In contrast, in the fatigued condition, a moderate negative correlation was detected between changes in DFAα1 from the warm-up to cool-down and total work, indicating that participants who performed greater amounts of work tended to exhibit larger reductions in DFAα1 following 20-min of intense exercise (Figure 6).

**Figure 6.**
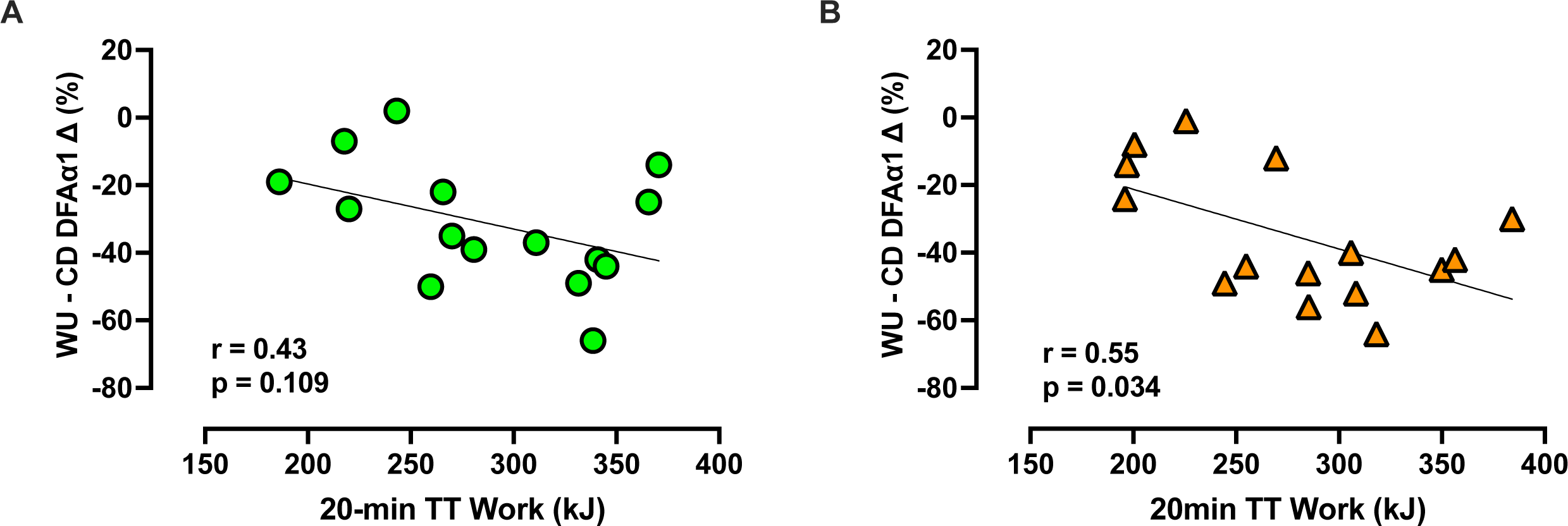
Relationships between changes in DFAα1 from the warm-up (WU) to cool-down (CD) and total work performed during the 20min TT in rested (Panel A) and fatigued (Panel B) conditions. Regression lines, Pearson correlation coefficients, and p values are provided for each panel. n=15 for both panels.

## Discussion

Our findings reinforce the sensitivity of DFAα1 to both exercise intensity and acute training stress while also demonstrating its potential to assess readiness-to-train and durability. As expected, DFAα1 declined progressively with increasing exercise intensity during both the warm-up and cool-down, with significantly lower values following 20 min of intense exercise; however, in contrast to our hypothesis, DFAα1 responses during the warm-up, 20-min TT, and cool-down, did not differ significantly between the rested and fatigued conditions. This may be due to the marginal, albeit significant, reduction (3.4%) in fatigued 20-min TT performance compared to rested performance. Consequently, any potential effect of the fatiguing protocol on DFAα1 may have been undetectable—likely falling within the typical day-to-day variability of the measurement (Van Hooren, Bongers, et al., 2023). Despite these limitations, across the two TTs, decreases in pre-TT DFAα1 values (i.e., warm-up DFAα1) were associated with reductions in performance (Figure 4), supporting the utility of evaluating between-session changes in DFAα1 to identify changes in day-to-day training readiness and performance.

Within-session deviations in DFAα1 responses during or following exercise performance trials may signal the onset and magnitude of physiological deterioration from prolonged exercise, offering a potential marker of exercise durability. Notably, we observed a decline in DFAα1 between the 10- and 20-min time points of the 20-min TT (Figure 3), indicating a progressive loss of signal correlation with sustained high-intensity efforts. In addition, post-TT DFAα1 values (i.e., cool-down) were significantly lower than pre-TT warm-up values across all relative intensities, regardless of condition (Figure 2), suggesting increased systemic perturbation and fatigue-induced physiological stress. Importantly, individuals with higher fitness levels (i.e., greater W·kg^−1^) exhibited lower average DFAα1 values during the 20-min TT (Figure 5). This pattern may indicate that fitter individuals are capable of tolerating and sustaining higher levels of internal load—potentially a signature of enhanced exercise durability. In the fatigued 20-min TT, participants who completed more total work also exhibited greater reductions in DFAα1 from warm-up to cool-down (Figure 6). Together, these results support the utility of DFAα1 as a sensitive marker of relative physiological strain during exercise and underscore its potential as a dynamic, non-invasive tool for monitoring in-session physiological status and individual resilience to exercise stress.

### DFAα1 - Readiness-to-Train

Between-session deviations in DFAα1 measurements at fixed external loads (e.g., cycling power output [PO]) may signal residual physiological strain from previous exercise to assess an athletes’ “readiness-to-train.” DFAα1 measurements taken during low-intensity exercise (i.e., 90% of the first ventilatory threshold [VT_1_]) can be used to differentiate between physiological states following light and heavy exercise training doses within short recovery windows (Schaffarczyk et al., 2022). In alignment with these findings, DFAα1 measurements during the warm-up and cool-down were, on average, 3.4 % and 8.0 % lower in the fatigued condition, respectively—although differences between conditions were not significant (Figure 2). Notably, DFAα1 reliability research indicates that the smallest worthwhile change (SWC) for DFAα1 is 5.5% in rested conditions and 8.0% in fatigued conditions (Van Hooren, Bongers, et al., 2023). As the observed reductions fell within this range of typical day-to-day variability, and, given the marginal reduction in 20-min TT performance, it remains unclear whether DFAα1 can reliably distinguish between low and high training loads. Additionally, there was no significant between-session difference in DFAα1 responses during the 20-min TT, even after considering differences in PO between the rested and fatigued trials (Figure 3). Accordingly, a more intensive fatigue protocol may be necessary to elicit larger reductions in performance to determine whether DFAα1 can effectively detect between-session variations in training load.

Despite the absence of distinct between-session effects, we observed that decreases in DFAα1 values during the warm-up were moderately associated with reduced 20-min TT performance, independent of training load condition (Figure 4).This finding supports the utility of DFAα1 as a readiness-to-train marker and aligns with previous research suggesting that submaximal DFAα1 measurements taken during standardized warm-ups may reflect acute physiological status (Schaffarczyk et al., 2022). Practically, a lower pre-training DFAα1 compared to an athlete’s typical baseline could serve as an indicator of a reduced capacity to tolerate a high training stimulus, signaling the requirement to adjust training load for an upcoming session (Figure 4).

### DFAα1 – Durability

Several in-session observations in the current study suggest that DFAα1 may serve as a marker for the onset and magnitude of physiological deterioration from prolonged exercise, providing insights into exercise durability. For example, we observed a 11% and 24% decline in DFAα1 between the 10- and 20-min during the rested and fatigued conditions, respectively (Figure 3)—although these findings were not different between conditions. Additionally, post-TT DFAα1 values were significantly lower at each relative intensity during the CD compared to the WU, regardless of condition (Figure 2). Together, these findings provide evidence that DFAα1 can detect increases in physiological perturbation, both during and following high-intensity exercise. Practically, comparing DFAα1 at fixed work rates—possibly throughout an extended training session—to known DFAα1-work rate responses established at baseline, may provide a valuable method to quantify physiological shifts from intense exercise and indicate altered states of exercise durability.

In alignment with these findings, we observed that the change in DFAα1 from the warm-up to cool-down was negatively associated with total work performed during the 20-min TT, albeit only in the fatigued condition (Figure 6). This finding is similar to previous research showing greater declines in DFAα1 following 40-min of fixed-intensity exercise compared to 20-min (Ajayi et al., 2025). These results suggest that athletes capable of sustaining higher workloads under fatigue also exhibit greater physiological disruption, and that DFAα1 decoupling from baseline responses may reflect the accumulated stress of an exercise session. Similar investigations have identified HR and breathing frequency as additional non-invasive indicators of exercise durability, demonstrating significant decoupling from baseline values after 2.5 hours of moderate intensity cycling (Rothschild et al., 2025). Given that DFAα1, HR, and breathing frequency can all be monitored using wearable sensors (Gronwald et al., 2025), the combined assessment of these metrics may offer practical tools for endurance athletes and coaches to detect reductions in exercise durability in real-world training settings.

Participants with higher fitness levels (i.e., W·kg^-1^) tended to exhibit lower DFAα1 values during their 20-min TTs (Figure 5). As lower DFAα1 values are thought to reflect higher degrees of systematic dysregulation from exercise stimuli (Rogers & Gronwald, 2022; Schaffarczyk et al., 2022), fitter individuals may better tolerate and sustain higher internal loads (i.e., superior durability). Previous investigations have also identified relationships between fitness and DFAα1 response, with lower sustained DFAα1 values reported in fitter participants during 10 km runs (Gronwald et al., 2020), suggesting that both fitness and durability may modulate exercising DFAα1 responses.

### Experimental Considerations

Our results provide further evidence of DFAα1’s sensitivity to exercise intensity and physiological stress aligning with prior reports showing an attenuation in DFAα1 during prolonged exercise near the maximal lactate steady state (van Rassel et al., 2024), relative to VO_2_max (Casties et al., 2006), and in response to prolonged or incremental endurance efforts (Gronwald et al., 2021; Rogers, Mourot, et al., 2021; Van Hooren, Mennen, et al., 2023). Despite these collective findings, it is important to note that many of these observations are based on group-averaged data. Interestingly, several individual DFAα1 responses did not align with that of the group, with some participants showing no substantial changes in DFAα1 following the 20min TT and large inter-individual variability in DFAα1 during the warm-up and cool-down—even when exercise intensity was scaled relative to each participant’s FTP (Figure 2). These findings highlight considerable individual differences in DFAα1 responses to exercise and suggest that group-level trends may not fully capture the complexity of individual physiological adaptations or stress responses. Indeed, prior work has demonstrated that agreement between HRV-derived thresholds and ventilatory thresholds improves when analyses considers each participants DFAα1 response to incremental exercise (Rogers et al., 2024), highlighting the importance of individualized interpretation. Future research should aim to explore these inter-individual differences in greater detail, with a focus on identifying the underlying physiological mechanisms driving this variability and evaluating the feasibility of personalized DFAα1-based monitoring strategies in both research and applied settings.

This investigation included remotely collected data from 5 of the 19 participants to highlight the feasibility and practical advantages of our purposed techniques. Remote-based research applications allows training to be prescribed and monitored outside of laboratory settings, enhancing accessibility and real-world applicability of DFAα1 (Andriolo et al., 2024). By utilizing commercially available heart rate monitors and technologies, DFAα1 has the potential to provide real-time physiological feedback, supporting a more individualized approach to endurance training. While factors like sensor placement, calibration, and unsupervised workouts introduce variability, remote DFAα1 monitoring remains a promising tool for assessing endurance adaptations, athlete readiness-to-train, and exercise durability, in real-world settings (Andriolo et al., 2024).

## Conclusion

This study highlights DFAα1’s sensitivity to acute exercise intensity and stress, reinforcing its potential as a physiological marker reflecting the dynamic interplay between internal and external workload (Michael et al., 2017; Rogers & Gronwald, 2022; White & Raven, 2014). In this context, our study demonstrates that DFAα1 tracks both acute intensity-related perturbations and accumulated fatigue, with measurable associations to performance outcomes and fitness levels supporting its utility as a readiness-to-train marker when measured pre-exercise, and as an index of exercise durability when tracked throughout or following prolonged efforts. Together, our findings support the utility of DFAα1 as a non-invasive, real-time tool for training monitoring, offering valuable guidance for adjusting training loads and enhancing endurance performance through more precise internal load management.

## Practical implications

- Monitoring DFAα1 during a warm-up may help athletes and coaches identify whether they are prepared for intense training or competition, with lower values potentially signalling a need for more recovery, reduced training intensities, or training volumes.
- Changes in DFAα1 during and after exercise may reflect how well an athlete handles sustained physical stress—offering a potential tool to assess exercise durability.
- Lower DFAα1 values during intense cycling efforts may indicate enhanced physiological readiness and an enhanced capacity to cope with prolonged physical stress

## Data Availability

Individual participant data are presented within the manuscript where appropriate. Additional supporting data are available from the corresponding author upon reasonable request.

## Acknowledgements

Funding: This investigation was supported by an operating grant from the Natural Sciences and Engineering Research Council of Canada (NSERC; grant number RGPIN-2025-06532) received by MJM. CVR was funded by NSERC, the NSERC CREATE Wearable Technology and Collaboration (We-TRAC) Training Program, an Alberta Innovates Graduate Student Scholarship for Data-Enabled Innovation, and an Alberta Graduate Excellence Scholarship. MR is the founder of a software application used for data processing, AI Endurance (https://aiendurance.com). All other authors declare no conflicts of interest. The authors would like to acknowledge the contributions of all participants, students, faculty, and staff, who assisted and made this investigation possible.

